# Molecular surveillance reveals widespread colonisation by carbapenemase and extended spectrum beta-lactamase producing organisms in neonatal units in Kenya and Nigeria

**DOI:** 10.1101/2022.01.06.22268735

**Authors:** Thomas Edwards, Christopher T Williams, Macrine Olwala, Pauline Andang’o, Walter Otieno, Grace N Nalwa, Abimbola Akindolire, Ana I Cubas-Atienzar, Toby Ross, Kemi Tongo, Emily R Adams, Helen Nabwera, Stephen Allen

## Abstract

**Objectives:** Neonatal sepsis, a major cause of death amongst infants in sub-Saharan Africa, is often gut derived. Impairments in immunity and the gut barrier in sick neonates allow colonisation by opportunistic pathogens such as *Enterobacteriaceae* to progress to blood stream infection. Colonisation by *Enterobacteriaceae* producing extended spectrum beta-lactamase (ESBL) or carbapenemase enzymes is particularly problematic and can lead to antimicrobial-resistant (AMR) or untreatable infections. We sought to explore the rates of colonisation by ESBL or carbapenemase producers and their genotypes in two neonatal units (NNUs) in West and East Africa.

**Methods:** Stool and rectal swab samples were taken at multiple timepoints from newborns admitted to the NNUs at the University College Hospital, Ibadan, Nigeria and the Jaramogi Oginga Odinga Teaching and Referral Hospital, Kisumu, western Kenya. Samples were tested for ESBL and carbapenemase genes using a previously validated qPCR assay with high resolution melt analysis. Kaplan-Meier survival analysis was used to examine colonisation rates at both sites.

**Results:** A total of 119 stool and rectal swab samples were taken from 42 infants admitted to the two NNUs. Six (14.3%) infants were extremely preterm (gestation <28 weeks), 19 (45.2%) were born by Caesarean section and 3 (8.6%) mothers were HIV positive. Median (IQR) duration of admission was 12.5 (5-26) days and 12 (28.6%) infants died. Overall, colonisation with ESBL (37 infants, 89%) was more common than with carbapenemase producers (26, 62.4%; P = 0.093). Median survival time before colonisation with ESBL organisms was 7 days and with carbapenemase producers 16 days (P=0.035). The majority of ESBL genes detected belonged to the CTX-M-1 (36/38; 95%), and CTX-M-9 (2/36; 5%) groups. The most prevalent carbapenemase was *bla*_NDM_ (27/29, 93%). Single *bla*_VIM_ (1/32, 3%) and *bla*_OXA-48_ genes (1/32, 3%) were also detected.

**Conclusions:** Gut colonisation of neonates by AMR organisms was common and occurred rapidly in NNUs in Kenya and Nigeria. Active surveillance of colonisation will improve the understanding of AMR in these settings and guide infection control and antibiotic prescribing practice to improve clinical outcomes.

**Highlights:**

- Colonisation with extended spectrum beta-lactamase (ESBL) or carbapenemase producing bacteria was common in two neonatal units in Kenya and Nigeria
- ESBL colonisation occurred in 89% of neonates, with a median colonisation time of 7 days
- Colonisation with carbapenemase producers occurred in 62% of neonates, with a median time to colonisation of 16 days
- The most common ESBL genes detected were of the CTX-M-1 family, whilst the most common carbapenemase detected was *bla*_NDM_

## Introduction

Sepsis is a major cause of neonatal morbidity and mortality, with an estimated 1.7 million cases globally in 2010, and 203,000 sepsis-attributable deaths (1). Neonatal sepsis has a higher incidence and mortality in sub-Saharan Africa (sSA) than in other regions (1, 2). It is often gut derived, with compromised immunity and an impaired gut barrier allowing colonisation by opportunistic pathogens such as *Enterobacteriaceae* to progress to blood stream infection (3). Preterm and low birth weight infants are at the greatest risk. Colonisation by antimicrobial resistant organisms (AROs) is particularly problematic and can lead to infections that are difficult or impossible to treat.

Infections with extended spectrum beta-lactamases (ESBL)-producing *Enterobacteriaceae* or carbapenemase producing organisms (CPOs), which confer resistance to 3^rd^ generation cephalosporins and carbapenems respectively, are associated with poor clinical outcomes due to the increased likelihood of treatment failure (4). Globally, organisms that produce either ESBL or carbapenemase enzymes, particularly *Klebsiella pneumoniae*, are a common cause of outbreaks in neonatal units (NNU) (5-7), often due to horizontal transmission from a colonised infant admitted to the ward (8). Mortality rates of up to 64% have been reported (9), driven by the lack of effective treatment options. Outbreaks of ESBL/CPO *K. pneumoniae* have been described frequently in African neonatal units (10) caused by contaminated intravenous fluids/antibiotics (11), or associated with gut colonisation (12). Treatment options for neonatal infections with this resistance profile in sub-Saharan Africa (sSA) are severely restricted, due to a lack of alternative antibiotics and contraindications in this age group (10, 13).

Data on the prevalence of gut colonisation in NNUs in sSA is limited due to a lack of routine surveillance. Defining ARO epidemiology and colonisation rates in NNUs in sSA is critical for guiding antimicrobial stewardship and to assess the impact of interventions (14). Typically, culture-based methods from rectal swabs or stool samples (15) are used for detecting colonisation. However, increasingly, molecular tools are used to screen samples directly for the presence of AMR genes, foregoing a culture step, providing faster results and increased throughput (16, 17).

We sought to explore the frequency and rates of gut colonisation by AROs in two NNU’s, one in West and one in East Africa, using highly multiplexed molecular diagnostics.

## Methods

### Study sites

The study was facilitated by the <u>Neonatal Nutrition Network for sSA</u> established in 2018 with the goal of building research capacity and establishing an environment for future trials of neonatal nutritional and other interventions (18). Of the 7 secondary/tertiary NNUs in Nigeria and Kenya engaged in the network, the study sites were the NNUs at the University College Hospital, Ibadan, Nigeria and the Jaramogi Oginga Odinga Teaching and Referral Hospital (JOOTRH), Kisumu, western Kenya.

Over a 6-month period between August 2018 and May 2019, the Ibadan NNU admitted 488 babies aged less than 48 hours of which 94/444 (21.2%) were very low birth weight (<1.5 kg) and 117/482 (24.3%) were very preterm. The Kisumu NNU admitted 584 babies aged less than 48 hours of which 100/579 (17.3%) were very low birth weight (<1500g) and 88/550 (16.0%) were very preterm (gestation 28-32 weeks).

### Ethics

The study was approved by the Liverpool School of Tropical Medicine Research Ethics Committee (study number 18-021), the University of Ibadan/University College Hospital Ethics Committee (UI/EC/18/0446) and the JOOTRH Ethics Review Committee (ERC.IB/VOL.1/510).

### Participants and data collection

Parents of neonates admitted to the NNUs were provided with information about the study and asked to provide informed consent. Clinical details of enrolled neonates were extracted from case records and entered into a case report form which was updated with patient outcomes upon discharge from the unit. Suspected sepsis was based on clinical assessment that advised starting or changing antibiotic treatment. Confirmation of sepsis by laboratory analysis was rarely available. Antibiotic usage data for each participant was also available from the Ibadan NNU.

### Sampling

Samples of neonatal stool were taken by swabbing faecal material from diapers or, if no stool was present, a rectal swab. Swabs were immediately frozen at -20° C prior to DNA extraction. Sampling was scheduled to occur on a weekly basis for six weeks post-admission. However, in practice, samples were often taken at different timepoints due to the high clinical workload for nursing staff.

### DNA extraction and molecular detection of AMR markers

DNA was extracted from the stool samples and rectal swabs using a Qiagen Fast DNA stool mini kit (Qiagen, Germany), following the manufacturer’s instructions. DNA was shipped to the UK for molecular analysis. Antimicrobial resistance genes were detected using a previously validated in-house high resolution melt (HRM) analysis qPCR assay (19) that detects the five main carbapenemase genes (*bla*_VIM_, *bla*_IMP-1_, *bla*_KPC_, *bla*_NDM_, *bla*_OXA-48_) and ESBL genes (CTX-M groups 1 and 9). Briefly, the 12.5μl reactions contained 6.25μl 2x Type-It HRM mix (Qiagen, Germany), 1μl primer mix (final primer concentrations range from 100nM to 500nM), 2.75μl molecular grade water and 2.5μl DNA. Reactions were carried out using a Rotor-Gene Q (Qiagen, Germany), with end point detection carried out via HRM. Positive controls were DNA samples extracted from isolates carrying each AMR gene as detailed previously (19).

### Statistical analysis

A patient was defined as CPO or ESBL colonised on the date that a carbapenemase or ESBL gene was detected by the HRM analysis qPCR assay. Patients were assumed not to be colonised until a positive sample was obtained. Rates of colonisation of both CPO and ESBL producers were described using Kaplan-Meier survival curves (GraphPad Prism), with patient’s data censored at the point of their last sample (15).

To calculate the proportion of the various AMR genes for both the ESBL and CPO genotypes in the cohort, all genes detected in an individual patients’ samples across all time points were included and collated. Genes that were isolated in multiple samples in a single patient were only counted once and when first identified, as these were assumed to belong to the same colonising organism. Associations between clinical variables and ARO colonisation were analysed with Chi-Squared and Fishers Exact Tests using GraphPad Prism Version 5 (GraphPad Software Inc, United States). A p-value <0·05 was considered significant.

## Results

### Demographics and clinical characteristics

A total of 42 neonates were enrolled in the study, including 24 in Ibadan and 18 in Kisumu (Table 1). Overall, 24 (57.1%) were female, and 6 (14.3%) were extremely preterm (gestational age <28 weeks). Suspected sepsis based on clinical assessment occurred in the majority of infants (30/42; 71.4%).

**Table 1.** Characteristics of the study participants, colonisation and mortality

| Characteristic | Ibadan<br>n (%) | Kisumu<br>n (%) | Total<br>n (%) |
| --- | --- | --- | --- |
| Participants | 24 (57.1) | 18 (42.9) | 42<br>(100.0) |
| Female | 14 (58.3) | 10 (55.6) | 24 (57.1) |
| Very low birthweight (<1500g) | 24 (100) | 4 (22.2) | 28 (66.7) |
| Very preterm (gestational age <28 weeks) | 1 (4.2) | 5 (27.8) | 6 (14.3) |
| Caesarean section delivery | 13 (54.2) | 6 (33.3) | 19 (45.2) |
| Mother HIV positive | 0 (0.0) | 3 (16.7) | 3 (8.6) |
| 1 or more episodes of suspected sepsis | 12 (50) | 18 (100.0) | 30 (71.4) |
| 1 or more episodes of necrotising enterocolitis | 5 (20.8) | 1 (5.5) | 6 (14.3) |
| ESBL colonised<sup>1</sup> | 22 (91.6) | 14 (77.8) | 36 (85.7) |
| CPO colonised<sup>1</sup> | 20 (83.3) | 8 (44.4) | 28 (66.7) |
| Uncolonized by antimicrobial-resistant organism<sup>1</sup> | 0 (0.0) | 2 (11.1) | 2 (4.8) |
| Mortality in neonatal unit | 5 (20.8) | 7 (38.9) | 12 (28.6) |

Notes:
ESBL = extended spectrum beta-lactamase
CPO = carbapenemase producing organism
<sup>1</sup>At any time during admission

### Sampling

A total of 58 samples were obtained from the 24 neonates in Ibadan between 1- and 56-days post admission. The average number of infants sampled per day between days 1-15, 16-30 and 31-45 were 1.9, 1.3 and 0.4, respectively. In Kisumu, 57 samples were obtained from the 18 neonates (Fig.S1), taken between 1- and 46-days post admission. The average number of infants sampled per day between days 1-15, 16-30 and 31-45 were 2.1, 1.5 and 0.5, respectively.

### Antibiotic usage

Antibiotic usage data was only available for the neonates in Ibadan NNU; antibiotics were used for both prophylaxis in newborns with risk factors for infection and treatment of suspected sepsis. The antibiotics received by the cohort were amikacin (received by all neonates), ampicillin/sulbactam (95.2%), ceftazidime (42.9%), gentamycin (38.1%), levofloxacin (19.1%), metronidazole (14.3%), piperacillin/tazobactam (9.52%), clindamycin (9.52%), cefotaxime (4.8%) ciprofloxacin (4.8%), vancomycin (4.8%), and meropenem (4.8%).

### Colonisation

The overall colonisation rates among the study participants were 88.0% for ESBL and 64.3% for CPO organisms. A total of 50.0% (9/18), 5.5% (1/18), and 33.3% (6/18) of neonates in the Kisumu NNU were colonised by ESBL, CPO, or both ESBL/CPO, respectively at any point during their admission (Fig.1A.). Colonisation rates were higher in Ibadan, with 79.2% (19/24) of neonates colonised by both ESBL/CPO organisms during their admission, and only 4.2% (1/24) uncolonised by either organism. Of the three infants who remained uncolonised across the two sites, all submitted a single sample on days one or two, with one being discharged from the ward and the remaining dying during the study. Colonisation according the demographic and clinical variables is shown in supplementary table 1.

**Fig 1.**
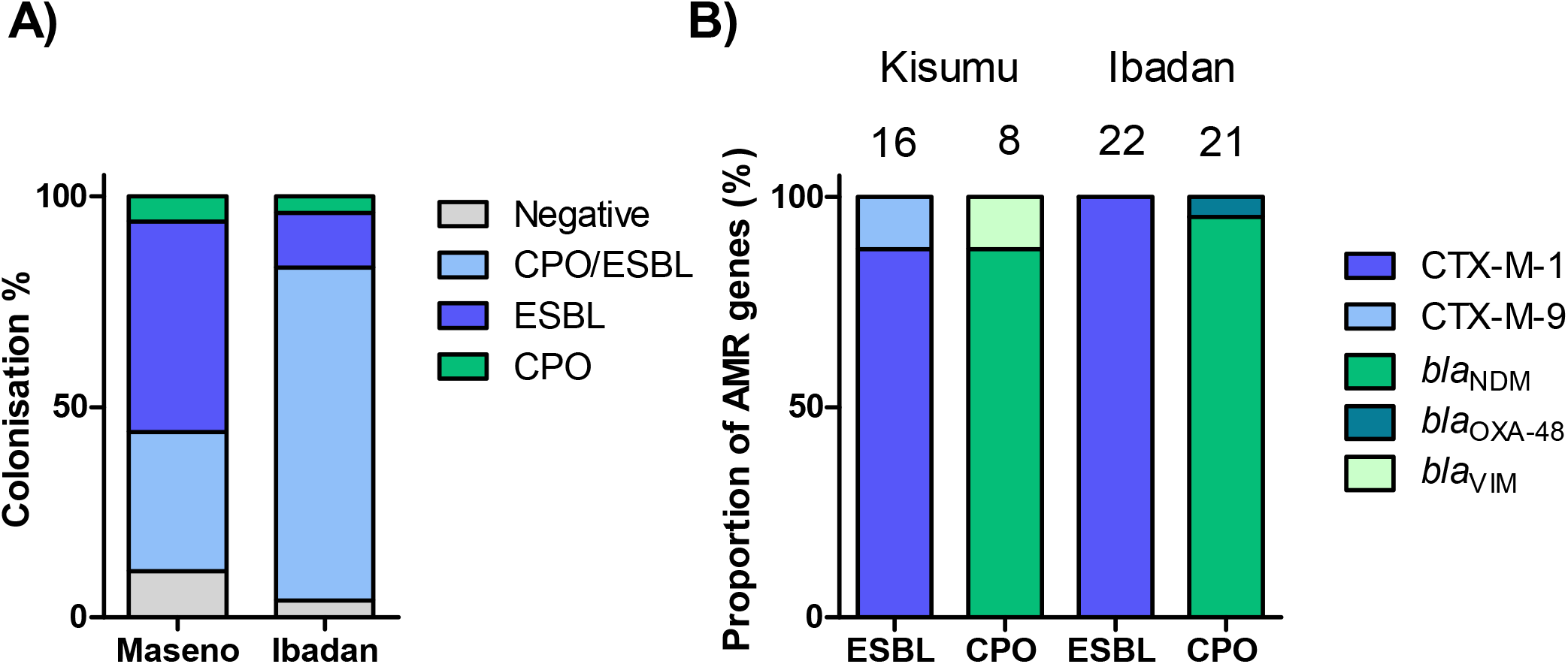
**A)** The percentage of participants at each site colonised by ESBL, CPO or both during the study period. **B)** Number of participants at each site positive for various ESBL and carbapenemase genes identified during the study.

### Colonisation rates

Longitudinal sampling revealed that colonisation occurred rapidly (Fig.2) and had often occurred by the first sampling point (35/42; 83.3%). Median survival time to colonisation by ESBL organisms was 7 days with a maximum of 97% of colonisation by day 45 (Fig.3). In contrast, the rate of CPO colonisation was significantly lower (P=0.035, Mantel-Cox Test), with a median survival time before colonisation of day 16, but also with a maximum of 97% by day 45. Colonisation was also dynamic; in 13 (31%) cases either ESBL or CPO carriage was detected but then absent in a subsequent sample.

**Fig 2.**
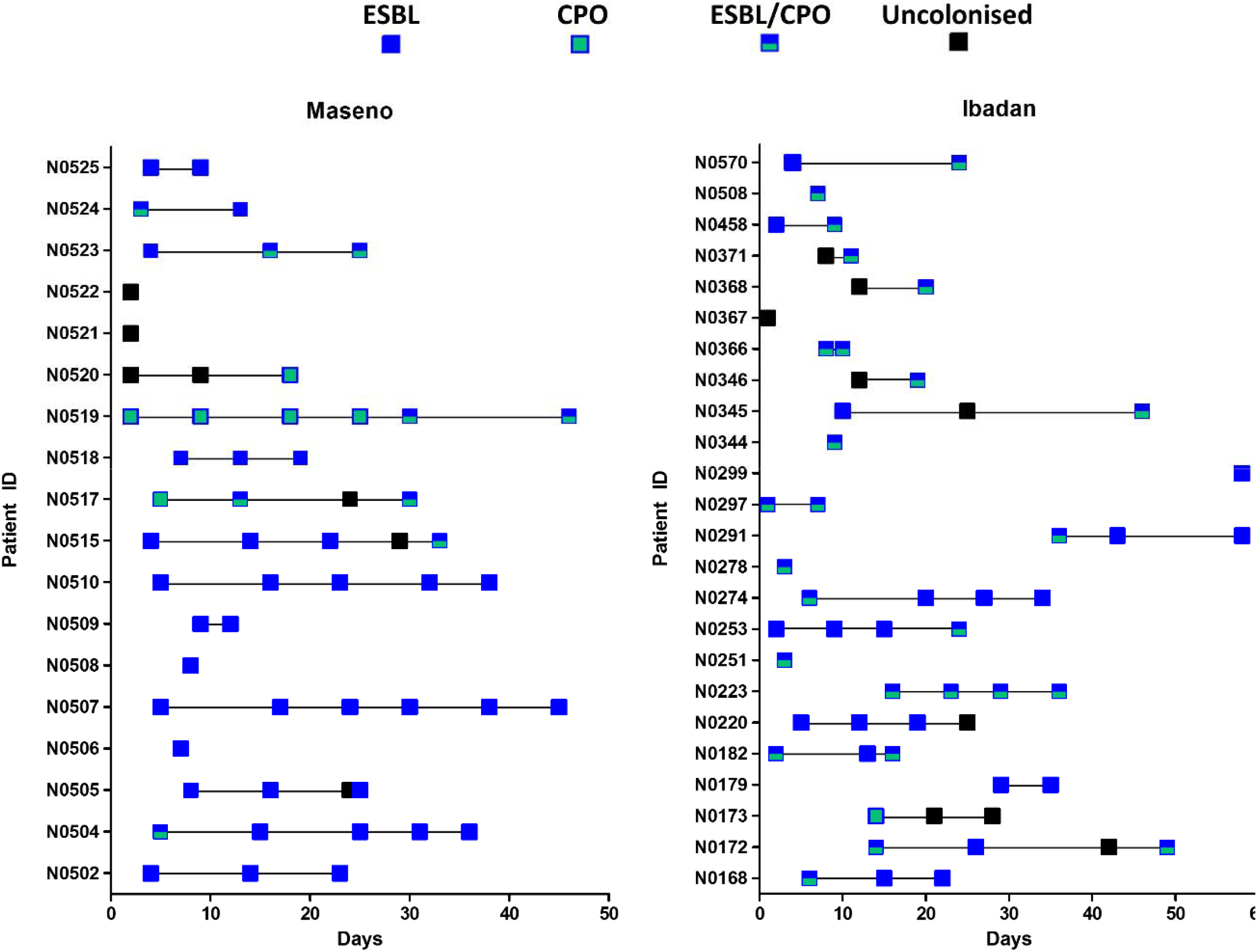
Longitudinal acquisition, carriage and loss of ESBL and CPOs according to study site. Note: Each box represents a stool or rectal swab sample. Days are counted from the admission of each individual participant and do not run concurrently.

**Fig 3.**
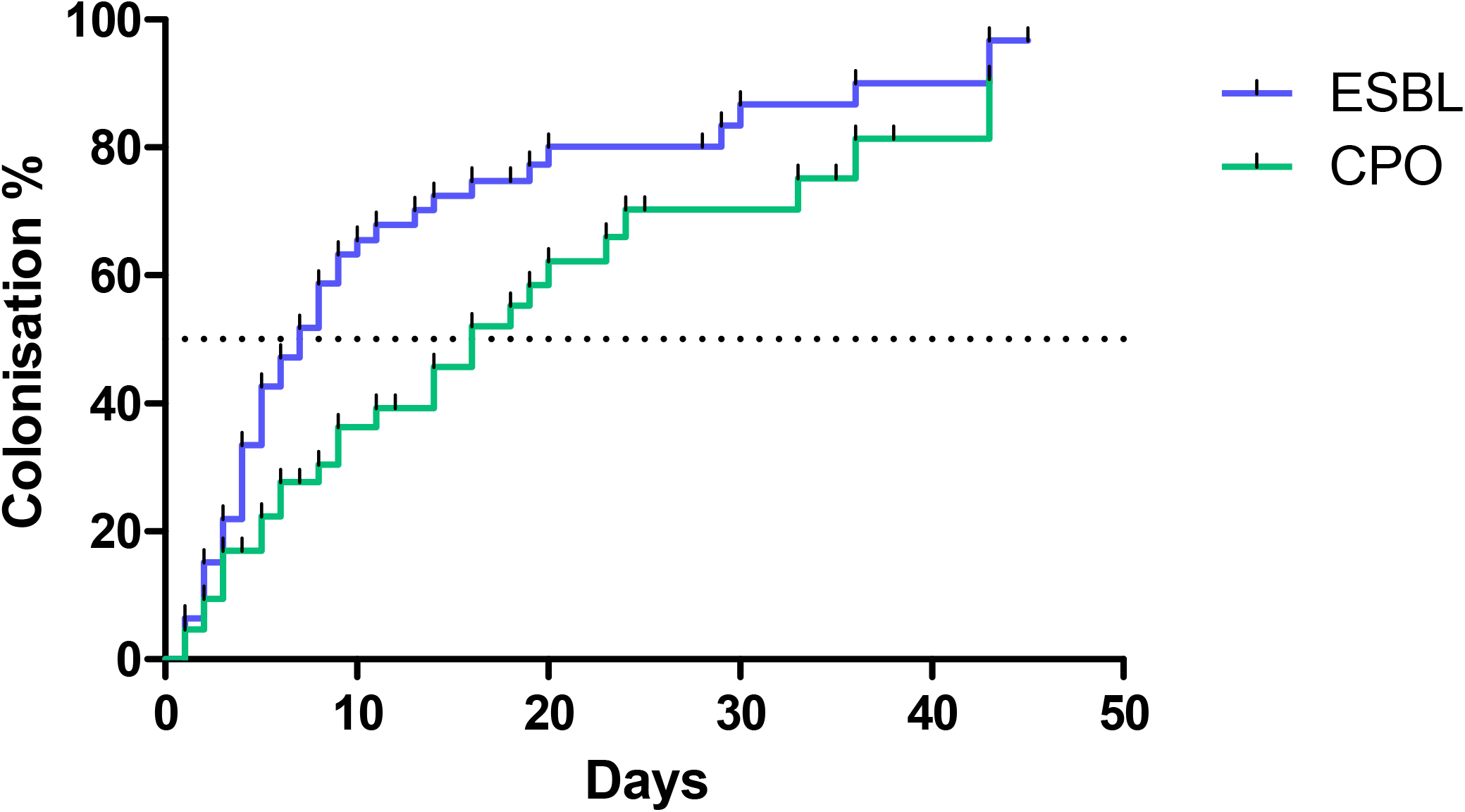
Kaplan-Meier survival analysis showing time to first colonisation by ESBL and CPOs Notes: Dotted line indicates time to 50% of infants colonised. Notes: Dotted line indicates time to 50% of infants colonised. ESBL = extended spectrum beta-lactamase CPO = carbapenemase producing organism

### Molecular epidemiology of AMR genes

In Kisumu, the majority of ESBL genes detected belonged to the CTX-M-1 family (14/16 87.5.%), and CTX-M-9 (2/16 12.5%) (Fig.1B.). The most prevalent carbapenemase was *bla*_NDM_ (7/8, 87.5%). A single *bla*_VIM_ gene was also detected (1/8, 12.5%). In Ibadan, CTX-M-1 was again the dominant ESBL family (22/22, 100%), and *bla*_NDM_ the dominant carbapenemase 20/21 (95.2%). One sample was positive for *bla*_OXA-48_ (1/22, 4.8%). We did not detect any of the carbapenemases *bla*_KPC_ or *bla*_IMP_genes in either site.

## Discussion

Using a combination of longitudinal sampling and molecular testing for a panel of AMR genes we were able to demonstrate rapid rates of colonisation of neonates admitted to two neonatal units by ESBL and carbapenemase producing organisms. The prevalence of colonisation by ESBLs in infants admitted to both the Kisumu and Ibadan NNUs was higher than previously reported in sSA NNUs including Tanzania (12) and Kenya (20). A systematic review of gut ESBL colonisation in sSA reported rates ranging from 25% to 74% (21), which are all below the levels found in our study. Colonisation of infants with ESBL producers was rapid, with mean time to colonisation of 8 days. This is comparable to studies in Asia (22) where the use of carbapenems and 3 ^rd^ generation cephalosporins is widespread. In a minority of infants, colonisation was dynamic, with acquisition and loss of either ESBL or CPO bacteria occurring during the study. Whilst colonisation can last for months or even years (23), rapid acquisition and loss has been shown in travellers to areas with high ESBL prevalence (24).

Data on gut colonisation by CPOs in sSA is limited; however, this is assumed to be less common than ESBL colonisation, in part due to the lower usage of expensive carbapenem drugs in this setting (25). A recent study from Nigeria, including patient isolates from the same hospital involved in our study, reported 69.1% of a cohort of hospital inpatients were colonised by CPOs, and highlighted the circulation of *bla*_NDM_ and *bla*_VIM_ carbapenemases (26). A country specific meta-analysis found the prevalence of carbapenem resistance amongst *Enterobacteriaceae* to be 5% in Nigeria and 0.3% in Kenya (27). The proportion of neonates colonised by CPOs in Ibadan exceeds the highest proportions reported in previous meta-analysis studies (28) carried out in non-NNU hospital and community settings in Africa and highlights that NNUs could be a major reservoir of CPOs.

The majority of CTX-M ESBL genes detected were in CTX-M group one, which includes *bla*_CTX-M-15_ and *bla*_CTX-M-1_ enzymes (29). These are the most widespread ESBL genes globally and have previously been reported as the most prevalent genes in sSA (29). The discovery of *bla*_NDM_ as the most common carbapenemase gene present in both the Ibadan and Kisumu tertiary NNUs is in agreement with previous studies; A 2012-2014 survey found *bla*_NDM_ and *bla*_VIM_ genes to be the only reported carbapenemases in both Nigeria and Kenya (30). A high prevalence of *bla*_NDM-1_ was found in the Ibadan NNU despite only a single dose of carbapenem antibiotics being given over the study period. However, 95.2% of neonates in Ibadan received a β-lactam antibiotic, and 46.7% a 3rd generation cephalosporin, and the use of these antibiotics could potentially select for bacteria producing any circulating enzyme capable of providing resistance, including carbapenemases. The frequent co-localisation of *bla*_NDM-1_ with other AMR genes on mobile genetic elements means that co-selection of *bla*_NDM-1_ can also occur during exposure to other antibiotic classes such as aminoglycosides (31).

The transmission of AROs from the hospital environment to the neonatal gut has been well documented, with health care workers (32), sinks and taps (33), surface environments (34) and maternal colonisation (32) all implicated as potential transmission sources. Molecular surveillance coupled with immediate patient isolation has been demonstrated to be an effective infection control measure in intensive care units (35), and the enhanced speed of molecular testing can be useful in ensuring prompt intervention to prevent hospital or unit wide outbreaks (15). However, these interventions require the infrastructure capacity for isolation units. Successful control measures that focus on preventing colonisation include the use of disposable gloves and gowns for single patient use (36), improved hand hygiene, and intensive bio-cleaning of wards (37). The assay we used in this study has since been validated in a dry-format, requiring only the addition of DNA and nuclease-free water to resuspend the dried reagents (38). This eliminates the need for a cold chain for shipping and storage, making it more applicable to this setting. By directly testing faecal samples or swabs without a culture step, this method can be used to provide same day results. Although this approach does not identify the bacterial species, there are companion molecular assays that can be run alongside if this information is desired (39).

Our study had several limitations. The small sample size limited our ability to explore associations between demographic and clinical variables and colonisation. Nevertheless, the very high proportions of ESBL and/or CPO colonised infants provides important epidemiological information to inform infection control strategies. Whilst utilising molecular methods has a number of advantages, including the identification of specific AMR genes, this approach prevented the linkage of AMR genes to particular isolates, and therefore, we could not determine the species or sequence type of the AROs detected. Our pragmatic approach to sampling around clinical care also meant that sampling did not occur at regular intervals, and not all participants were sampled within their first week of admission to the NNU.

This report highlights that gut colonisation of infants by AROs in NNUs in Kenya and Nigeria is common and occurs rapidly. The carbapenemase *bla*_NDM-1_ and group 1 *bla*_CTX-M_ genes were the most prevalent resistance genes at both sites. The study demonstrates that active surveillance of colonisation can be used to improve the understanding of AMR in NNUs in sSA, and these data could guide infection control and prescribing practice to improve clinical outcomes.

## Data Availability

All data produced in the present study are available upon reasonable request to the authors

## Author contributions

TE, KT, HN, ERA and SA conceptualised the study. PA, WO, MO, GN, AA, and KT managed the clinical studies, including collecting patient information and samples. TE, CTW, KT, HN, ERA and SA contributed to the experimental design and data analysis. TE, CTW and TR carried out laboratory experiments. TE, CW, AICA and SA wrote the first draft of the manuscript. All authors approved the final manuscript.

Acknowledgments

We acknowledge the technical staff in the diagnostic microbiology laboratories at Kenya Medical Research Institute, Centre for Global Health Research, Kisumu, Kenya and the University College Hospital, Ibadan, Nigeria. We thank the parents and NNU staff who very kindly supported this study.

## Additional information

Correspondence should be addressed to TE or SA.

---

**Supplementary table 1.** Occurrence of selected variables in participants according to colonisation with ESBL and CPOs

| Variable | ESBL [n (%)] |  | CPO [n (%)] |  |
| --- | --- | --- | --- | --- |
|  | Colonised<br>(n=36) | Uncolonised<br>(n=6) | Colonised<br>(n=28) | Uncolonised<br>(n=14) |
| Female | 18 (50.0) | 5 (83.3) | 17 (60.7) | 7 (50.0) |
| <28 weeks' gestation | 5 (13.9) | 0 (0.0) | 2 (7.1.) | 3 (21.4) |
| Mother HIV positive | 2 (5.6) | 1 (16.7) | 2 (7.1) | 1 (7.1) |
| C section | 13 (36.1) | 5 (83.3) | 15 (53.6) | 4 (28.6) |
| Suspected sepsis | 25 (69.4) | 5 (83.3) | 17 (60.7) | 13 (92.9) |
| NEC | 4 (11.1) | 2 (33.3) | 5 (17.9) | 1 (7.1) |
| Mortality in NNU | 9 (25.0) | 3 (50.0) | 6 (21.4) | 6 (42.9) |

Notes: <sup>1</sup>NEC; necrotising enterocolitis. <sup>2</sup>NNU; neonatal unit.
ESBL = extended spectrum beta-lactamase
CPO = carbapenemase producing organism

## Supplementary Figures

**Fig.S1.**
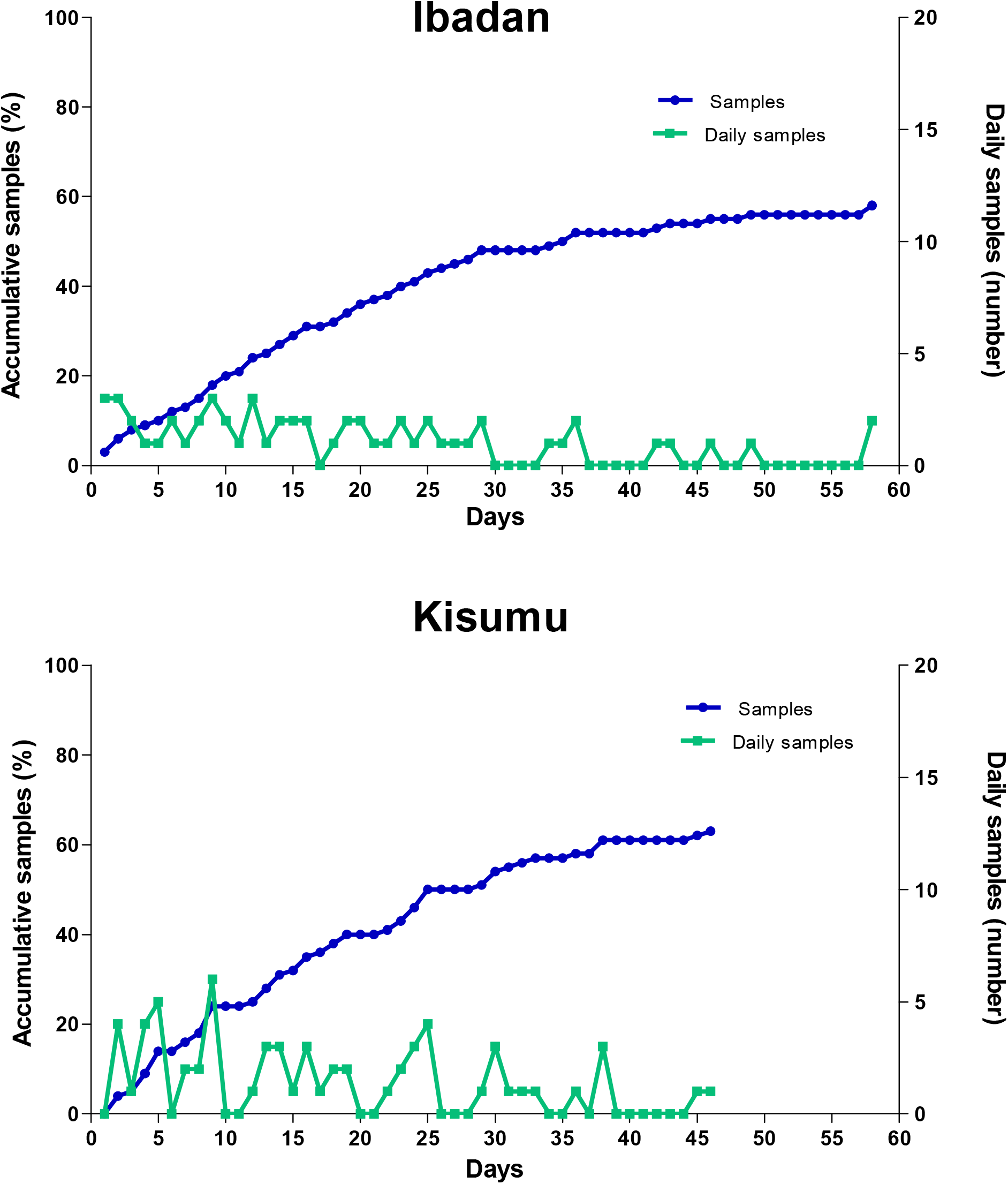
Sample collection rates in Kisumu and Ibadan NNUs. Days are counted from first day of life per participant

## References

1. Ranjeva SL, Warf BC, Schiff SJ. Economic burden of neonatal sepsis in sub-Saharan Africa. BMJ Glob Health. 2018;3(1):e000347.

2. Seale AC, Blencowe H, Manu AA, Nair H, Bahl R, Qazi SA, et al. Estimates of possible severe bacterial infection in neonates in sub-Saharan Africa, south Asia, and Latin America for 2012: a systematic review and meta-analysis. Lancet Infect Dis. 2014;14(8):731–41.

3. Haussner F, Chakraborty S, Halbgebauer R, Huber-Lang M. Challenge to the Intestinal Mucosa During Sepsis. Front Immunol. 2019;10(891).

4. Tumbarello M, Sanguinetti M, Montuori E, Trecarichi EM, Posteraro B, Fiori B, et al. Predictors of mortality in patients with bloodstream infections caused by extended-spectrum-beta-lactamase-producing Enterobacteriaceae: importance of inadequate initial antimicrobial treatment. Antimicrob Agents Chemother. 2007;51(6):1987–94.

5. Yin D, Zhang L, Wang A, He L, Cao Y, Hu F, et al. Clinical and molecular epidemiologic characteristics of carbapenem-resistant Klebsiella pneumoniae infection/colonization among neonates in China. J Hosp Infect. 2018;100(1):21–8.

6. Ghaith DM, Zafer MM, Said HM, Elanwary S, Elsaban S, Al-Agamy MH, et al. Genetic diversity of carbapenem-resistant Klebsiella Pneumoniae causing neonatal sepsis in intensive care unit, Cairo, Egypt. Eur. J. Clin. Microbiol. Infect. Dis.. 2019.

7. Chen D, Hu X, Chen F, Li H, Wang D, Li X, et al. Co-outbreak of multidrug resistance and a novel ST3006 Klebsiella pneumoniae in a neonatal intensive care unit: A retrospective study. Medicine. 2019;98(4):e14285.

8. Stapleton PJM, Murphy M, McCallion N, Brennan M, Cunney R, Drew RJ. Outbreaks of extended spectrum β-lactamase-producing Enterobacteriaceae in neonatal intensive care units: a systematic review. Arch Dis Child Fetal Neonatal Ed. 2016;101(1):72.

9. Stoesser N, Giess A, Batty EM, Sheppard AE, Walker AS, Wilson DJ, et al. Genome sequencing of an extended series of NDM-producing Klebsiella pneumoniae isolates from neonatal infections in a Nepali hospital characterizes the extent of community-versus hospital-associated transmission in an endemic setting. Antimicrob Agents Chemother. 2014;58(12):7347–57.

10. Cornick J, Musicha P, Peno C, Seager E, Iroh Tam P-Y, Bilima S, et al. Genomic investigation of a suspected Klebsiella pneumoniae outbreak in a neonatal care unit in sub-Saharan Africa. Microb Genom. 2021;7(11).

11. Okomo U, Senghore M, Darboe S, Bojang E, Zaman SMA, Hossain MJ, et al. Investigation of sequential outbreaks ofBurkholderia cepacia and multidrug-resistant extended spectrum β-lactamase producing Klebsiella species in a West African tertiary hospital neonatal unit: a retrospective genomic analysis. Lancet Microbe. 2020;1(3):e119–e29.

12. Marando R, Seni J, Mirambo MM, Falgenhauer L, Moremi N, Mushi MF, et al. Predictors of the extended-spectrum-beta lactamases producing Enterobacteriaceae neonatal sepsis at a tertiary hospital, Tanzania. Int. J. Med. Microbiol. Suppl. 2018;308(7):803–11.

13. Wen SCH, Ezure Y, Rolley L, Spurling G, Lau CL, Riaz S, et al. Gram-negative neonatal sepsis in low-and lower-middle-income countries and WHO empirical antibiotic recommendations: A systematic review and meta-analysis. PLoS medicine. 2021;18(9):e1003787.

14. Houghteling PD, Walker WA. Why is initial bacterial colonization of the intestine important to infants’ and children’s health? J Pediatr Gastroenterol Nutr. 2015;60(3):294–307.

15. Banerjee R, Humphries R. Clinical and laboratory considerations for the rapid detection of carbapenem-resistant Enterobacteriaceae. Virulence. 2017;8(4):427–39.

16. Ko YJ, Kim J, Kim H-N, Yoon S-Y, Lim CS, Lee CK. Diagnostic performance of the Xpert Carba-R assay for active surveillance of rectal carbapenemase-producing organisms in intensive care unit patients. Antimicrob Res Infect Control. 2019;8(1):127.

17. Williams CT, Edwards T, Adams ER, Feasey NA, Musicha P. ChloS-HRM, a novel assay to identify chloramphenicol-susceptible Escherichia coli and Klebsiella pneumoniae in Malawi. J Antimicrob Chemother. 2020. 74:5, 1212–1217

18. Nabwera HM, Wang D, Tongo OO, Andang’o PEA, Abdulkadir I, Ezeaka CV, et al. Burden of disease and risk factors for mortality amongst hospitalized newborns in Nigeria and Kenya. PLOS ONE. 2021;16(1):e0244109.

19. Edwards T, Williams C, Teethaisong Y, Sealey J, Sasaki S, Hobbs G, et al. A highly multiplexed melt-curve assay for detecting the most prevalent carbapenemase, ESBL and AmpC genes. Diagn Microbiol Infect Dis. 2020:115076.

20. Kagia N, Kosgei P, Ooko M, Wafula L, Mturi N, Anampiu K, et al. Carriage and Acquisition of Extended-spectrum β-Lactamase–producing Enterobacterales Among Neonates Admitted to Hospital in Kilifi, Kenya.Clin Infect Dis. 2019;69(5):751–9.

21. Lewis JM, Lester R, Garner P, Feasey NA. Gut mucosal colonisation with extended-spectrum beta-lactamase producing Enterobacteriaceae in sub-Saharan Africa: a systematic review and meta-analysis. Wellcome Open Res. 2019;4:160.

22. Roberts T, Limmathurotsakul D, Turner P, Day NPJ, Vandepitte WP, Cooper BS. Antimicrobial-resistant Gram-negative colonization in infants from a neonatal intensive care unit in Thailand. J Hosp. Infect. 2019;103(2):151–5.

23. Arcilla MS, van Hattem JM, Haverkate MR, Bootsma MCJ, van Genderen PJJ, Goorhuis A, et al. Import and spread of extended-spectrum beta-lactamase-producing Enterobacteriaceae by international travellers (COMBAT study): a prospective, multicentre cohort study. Lancet Infect Dis. 2017;17(1):78–85.

24. Kantele A, Kuenzli E, Dunn S, Dance D, Newton P, Davong V, et al. Real-time sampling of travelers shows intestinal colonization by multidrug-resistant bacteria to be a dynamic process with multiple transient acquisitions. bioRxiv. 2019:827915.

25. Musicha P, Cornick JE, Bar-Zeev N, French N, Masesa C, Denis B, et al. Trends in antimicrobial resistance in bloodstream infection isolates at a large urban hospital in Malawi (1998-2016): a surveillance study. Lancet Infect Dis. 2017;17(10):1042–52.

26. Ogbolu DO, Piddock LJV, Webber MA. Opening Pandora’s box: high level resistance to antibiotics of last resort in Gram negative bacteria from Nigeria. J Glob Antimicrob Res. 2020.21: 211–217

27. Mitgang EA, Hartley DM, Malchione MD, Koch M, Goodman JL. Review and mapping of carbapenem-resistant Enterobacteriaceae in Africa: Using diverse data to inform surveillance gaps. Int J Antimicrob Agents. 2018;52(3):372–84.

28. Manenzhe RI, Zar HJ, Nicol MP, Kaba M. The spread of carbapenemase-producing bacteria in Africa: a systematic review. J Antimicrob Chemother. 2014;70(1):23–40.

29. Bevan ER, Jones AM, Hawkey PM. Global epidemiology of CTX-M β-lactamases: temporal and geographical shifts in genotype. J Antimicrob Chemother. 2017;72(8):2145–55.

30. Kazmierczak KM, Rabine S, Hackel M, McLaughlin RE, Biedenbach DJ, Bouchillon SK, et al. Multiyear, Multinational Survey of the Incidence and Global Distribution of Metallo-β-Lactamase-Producing Enterobacteriaceae and Pseudomonas aeruginosa. Antimicrob Agents Chemother. 2015;60(2):1067–78.

31. Johnning A, Karami N, Tång Hallbck E, Müller V, Nyberg L, Buongermino Pereira M, et al. The resistomes of six carbapenem-resistant pathogens - a critical genotype-phenotype analysis. Microb Genom. 2018;4(11):e000233.

32. Tschudin-Sutter S, Frei R, Battegay M, Hoesli I, Widmer AF. Extended spectrum ß-lactamase-producing Escherichia coli in neonatal care unit. Emerg infect Dis. 2010;16(11):1758–60.

33. De Geyter D, Blommaert L, Verbraeken N, Sevenois M, Huyghens L, Martini H, et al. The sink as a potential source of transmission of carbapenemase-producing Enterobacteriaceae in the intensive care unit. Antimicrob Res Infect Control. 2017;6(1):24.

34. Bokulich NA, Mills DA, Underwood MA. Surface microbes in the neonatal intensive care unit: changes with routine cleaning and over time. J Clinical Microbiol. 2013;51(8):2617–24.

35. Zhou M, Kudinha T, Du B, Peng J, Ma X, Yang Y, et al. Active Surveillance of Carbapenemase-Producing Organisms (CPO) Colonization With Xpert Carba-R Assay Plus Positive Patient Isolation Proves to Be Effective in CPO Containment. Front. Cell. Infect. Microbiol.. 2019;9(162).

36. Harris AD, Morgan DJ, Pineles L, Magder L, O’Hara LM, Johnson JK. Acquisition of antibiotic-resistant Gram-negative bacteria in the Benefits of Universal Glove and Gown (BUGG) Cluster Randomized Trial. Clin Infect Dis. 2020.

37. Ferry A, Plaisant F, Ginevra C, Dumont Y, Grando J, Claris O, et al. Enterobacter cloacae colonisation and infection in a neonatal intensive care unit: retrospective investigation of preventive measures implemented after a multiclonal outbreak. BMC Infect Dis. 2020;20(1):682-.

38. Cubas-Atienzar AI, Williams CT, Karkey A, Dongol S, Sulochana M, Rajendra S, et al. A novel air-dried multiplex high-resolution melt assay for the detection of extended-spectrum β-lactamase and carbapenemase genes. J Glob Antimicrob Res. 2021;27:123–31.

39. Edwards T, Sasaki S, Williams C, Hobbs G, Feasey NA, Evans K, et al. Speciation of common Gram-negative pathogens using a highly multiplexed high resolution melt curve assay. Sci Rep. 2018;8(1):1114.

